# Antibody Responses after SARS-CoV-2 Vaccination in Lymphoma

**DOI:** 10.1101/2021.06.05.21258311

**Authors:** Sean Hua Lim, Nicola Campbell, Marina Johnson, Debora Joseph-Pietras, Graham P Collins, Ann O’Callaghan, Christopher P Fox, Matthew Ahearne, Peter WM Johnson, David Goldblatt, Andrew J Davies

## Abstract

Individuals with lymphoid malignancies have an increased mortality risk from COVID-19. Paradoxically, this population is least likely to be protected by SARS-CoV-2 vaccination as a result of disease- or treatment-related immunosuppression. Current data on vaccine responses in persons with lymphoid malignancies is limited.

PROSECO is a UK multi-centre prospective observational study evaluating COVID-19 vaccine immune responses in individuals with lymphoma. This early interim analysis details the antibody responses to first- and second-SARS-CoV-2 vaccination with either BNT162b2 (Pfizer-BioNTech) and ChAdOx1 (AstraZeneca), in 129 participants. Responses are compared to those obtained in healthy volunteers.

The key findings of this interim analysis are first, 61% of participants who are vaccinated during or within 6 months of receiving systemic anti-lymphoma treatment, do not have detectable antibodies despite two doses of vaccine. Second, individuals with curable disease such has Hodgkin (6/6) and aggressive B-cell non-Hodgkin lymphoma (13/16) develop robust antibody levels to either first or second doses, when vaccinated > 6 months after treatment completion. Third, participants incurable, indolent lymphomas have reduced antibody levels to first and second vaccine doses, irrespective of treatment history. Finally, whilst there was no difference in antibody responses between BNT162b2 and ChAdOx1 in lymphoma participants, BNT162b2 induces 11-fold higher antibody responses than ChAdOx1 after the second dose in healthy donors.

These findings serve to reassure the community that individuals with treated Hodgkin and aggressive B-NHL can develop antibody responses to SARS-CoV-2 vaccine. Simultaneously it also highlights the critical need to identify an alternative strategy against COVID-19 for those undergoing systemic anti-lymphoma treatment, and for individuals with indolent lymphomas.

## Antibody Responses after SARS-CoV-2 Vaccination in Lymphoma

Individuals with lymphoid malignancies risk developing severe COVID-19 and are less likely to develop protective immune responses to SARS-CoV-2 vaccination because of disease- or treatment-related immunosuppression. Current data on vaccine responses in chronic lymphocytic leukaemia show antibody responses in 52-75% of individuals after the second dose.^1,2^ Vaccine responses after two doses in persons with other lymphoid malignancies remain undefined.

In this study (NCT04858568) we report antibody levels prior to vaccination, two weeks post-first dose and/or two-four weeks post-second dose in 129 participants with lymphoma.(table S1) Participants received either ChAdOx1 (AstraZeneca) or BNT162b2 (Pfizer-BioNTech) vaccines, ten-twelve weeks apart.^3,4^ IgG antibodies against SARS-CoV-2 Spike antigen were measured using an electrochemiluminescent assay (MesoScale Discovery®)^5^ and responses reported in Binding Antibody Units/ml (BAU/ml), calibrated against the WHO COVID-19 International Reference Serum. Antibody titres were compared to those in healthy volunteers.

Antibody titres were significantly reduced in participants “on treatment” (ongoing treatment or completed ≤ six months before vaccination) compared to “no treatment” (treatment-naive or completed therapy > six months before first vaccination).(figures 1A, S1) Post-second dose geomean titres (GMT) for both vaccines were 2·5 BAU/ml (95% CI 1·1-5·8) and 141·8 (75·6-266·0) for “on” vs. “no treatment”, respectively. There was no difference in antibody levels between the vaccines within lymphoma cases despite significant differences in healthy donors (second dose; ChAdOx1 206 (124-344); BNT162b2 2339 (1923-2844), p<0.0001).

**Figure 1.**
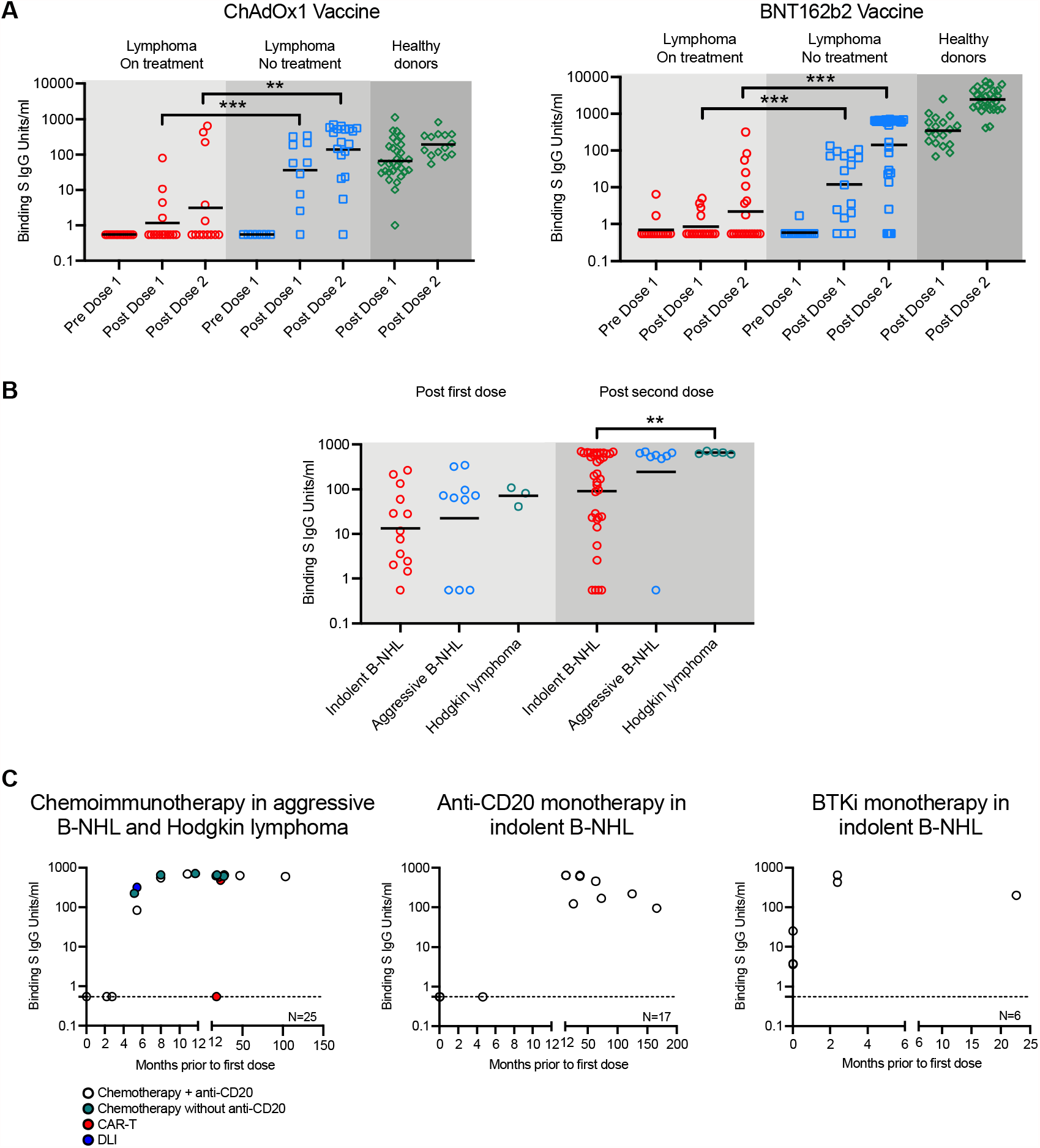
Anti-S IgG Response to First and Second Doses of SARS-Cov-2 Vaccination. **(A)** IgG antibodies against the SARS-CoV-2 S protein were measured using (MesoScale Discovery®)(MSD) and calibrated to the WHO International reference serum (NIBSC 20/136) prior to vaccination (pre dose 1), 4 weeks post-first vaccination (post dose 1) and/or 2-4 weeks post-second vaccination (post dose 2) with ChAdOx1 or BNT162b2 vaccines. Red circles represent participants with lymphoma in the “on treatment” group, blue squares represent participants with lymphoma in the “no treatment” group, and green triangles represent healthy donors. Participants “on treatment” were receiving systemic anti-lymphoma therapy at the time of administration of the first dose of vaccine, or completed treatment ≤6 months prior to the first vaccine dose, or commenced anti-systemic anti-lymphoma therapy <4 weeks after receiving the first vaccine dose. Participants with “no treatment” were treatment-naive or completed treatment >6 months prior to the first vaccine dose. (Median ages in years for ChAdOx1 (68;68) and BNT162b2 (70;70) in “on treatment” and “no treatment” groups, respectively; Healthy donors, median ages are 60 (range 23-70) and 43 (21-77) for ChAdOx1 and BNT162b2, respectively; No correlation was observed between anti-S levels and age for BNT162b2.) Binding Antibody Units per ml (BAU/ml) is shown. Anti-S IgG levels ≤ 0.55 BAU/ml are below the lower limit of detection. Geometric mean titres are shown. Significance was determine using two-tailed Mann-Whitney test, p<0.01(**), p<0.001(***). **(B)** Anti-S IgG levels in participants with lymphoma from the “no treatment” group, after first and second doses of both vaccines are shown. Red, blue and green circles represent indolent B-NHL, aggressive B-NHL and Hodgkin lymphoma, respectively. Geometric mean titres are shown. Significance was determine using two-tailed Mann-Whitney test, p<0.01(**). **(C)** Post-second vaccination anti-S IgG levels were compared to the timing of treatment completion. The X axes represent the number of months systemic anti-lymphoma treatment was completed prior to administration of the first vaccine dose. Participants who received their first dose of vaccine whilst on treatment are plotted at ‘0’ on the X axis. The graph on the left shows the number of months treatment had completed prior to administration of first dose of vaccine for participants with aggressive B-NHL and Hodgkin lymphoma. Types of treatment received were combination chemotherapy +/-anti-CD20, CAR-T cell therapy or donor lymphocyte infusions (DLI) post allogeneic stem cell transplantation. Similarly, the graphs in the middle and right show the number of months after completion of anti-CD20 monoclonal antibody therapy and Bruton Tyrosine Kinase inhibitor (BTKi, ibrutinib) monotherapy, respectively, in participants with indolent B-NHL.

Within the “no treatment” group, participants with curable disease (100% (6/6) Hodgkin and 81% (13/16) aggressive B-NHL) developed robust antibody levels after the first and second doses (Hodgkin: 652·2 (604·7-703·4); aggressive B-NHL:244·6 (31·12-1923)).(figure 1B) The exceptions were chimeric antigen receptor T-cell recipients where 3/3 and 1/2 had no detectable antibodies after the first and second dose, respectively. Reduced antibody levels were observed in indolent B-NHL (90·2 (40·15-202·4)). This is despite treatment-naivety or completion of treatment > three years previously. In participants who completed treatment prior to vaccination, those vaccinated ≤ six months of chemotherapy +/-anti-CD20, or anti-CD20 monotherapy, and ≤ two months of ibrutinib monotherapy had no detectable antibodies.(figure 1C)

In summary, individuals with curable lymphoma subtypes such as Hodgkin and aggressive B-NHL can develop robust serological responses as early as six months post-treatment. In contrast, individuals vaccinated whilst undergoing systemic anti-lymphoma therapy, and those with indolent lymphomas may have impaired serological responses and might benefit from further measures to protect them against COVID-19.

## Data Availability

The data from this study is available after study completion on request to the corresponding author.

## Contributors

SHL conceptualised, designed, supervised, wrote the original draft of the manuscript and contributed to the formal analysis. DG and MJ contributed to the methodology and formal analysis and verified underlying data. SHL, PJ, DG and AJD contributed to review and editing of the manuscript. SHL, NC and DJP were responsible for data curation and project administration. MA, CPF, GPC, AO, PJ, AJD and SHL recruited patients. SHL and AJD acquired funding. All authors had access to all the data reported in the study. The senior author had the final responsibility for the decision to submit for publication.

## Acknowledgements

The PROSECO study is funded by The Vaccine Blood Cancer Research Funding Consortium, and all charity partners including Blood Cancer UK (21009), and supported by Cancer Research UK Advanced Clinician Scientist Fellowship to SHL (A27179), Cancer Research UK/NIHR Southampton Experimental Cancer Medicine Centre funding and the NIHR Clinical Research Facility Southampton Research Biorepository (SRB). DG receives support from the NIHR Great Ormond Street Biomedical Research Centre. No pharmaceutical company contributed to this study or the writing of this article. This work is approved by local research ethics committees and the UK NHS Health Research Authority (IRAS 294739; 233768). Healthy controls were vaccinees who received vaccine as part of the government rollout and donated serum post vaccination for essay evaluation.

