## Supplementary material for "Antibody Responses after SARS-CoV-2 Vaccination in Lymphoma": Table S1, Figure S1

**Supplementary Information**

|  | **Hodgkin lymphoma** | **Aggressive B-NHL** | **Indolent B-NHL** | **Peripheral T/NK cell lymphoma** |
| --- | --- | --- | --- | --- |
| **Number of cases** | 12 | 34 | 79 | 4 |
| **Age in years,**  **median (range)** | 46 (23-79) | 67 (36-87) | 69 (24-93) | 63 (52-72) |
| **Gender** |  |  |  |  |
| **Male** | 10 | 18 | 50 | 3 |
| **Female** | 2 | 16 | 29 | 1 |
| **Vaccine type** |  |  |  |  |
| **BNT162b2** | 7 | 22 | 48 | 1 |
| **ChAdOx1** | 5 | 12 | 31 | 3 |
| **Disease subtypes**  **(no of cases)** | Not collected | DLBCL (26) | FL (34) | EATL (2) |
|  |  | Transformed FL (5) | CLL (17) | AITL (1) |
|  |  | EBV-related DLBCL (1) | MCL (10) | BIA-ALCL (1) |
|  |  | PCNSL (1) | LPL (8) |  |
|  |  | Double Hit (1) | MZL (7) |  |
|  |  |  | Hairy Cell Leukemia (1) |  |
|  |  |  | NLPHL (1) |  |
|  |  |  | Low grade B-NHL (1) |  |
| **Treatment status** |  |  |  |  |
| **None** | 0 | 0 | 20 | 0 |
| **Previous** | 6 | 17 | 30 | 2 |
| **Active** | 6 | 17 | 29 | 2 |
| **Number of lines of treatment,**  **median (range)** | 1 (1-4) | 3 (1-10) | 1 (1-7) | 2 |
| **Previous autologous stem cell**  **transplant** | 1 | 8 | 3 | 2 |
| **Previous allogeneic stem cell transplant** | 0 | 2 | 0 | 0 |
| **Remission status** |  |  |  |  |
| **CR/PR** | 12 | 24 | 48 | 4 |
| **PD** | 0 | 3 | 5 | 0 |
| **SD** | 0 | 0 | 18 | 0 |
| **Not yet assessed** | 0 | 7 | 7 | 0 |
| **Most recent therapy** | ABVD/AVD (5) | Acalabrutinib (2) | Ibrutinib (7) | Autograft (2) |
|  | ChlVPP/EVA/AVD (3) | Autograft (3) | Ibrutinib-venetoclax (1) | Cyclosporin (1) |
|  | BEACOPDac/AVD (2) | Bispecific antibody (1) | Obinutuzimab-CVP (4) | Prednisolone (1) |
|  | Nivolumab (1) | CAR-T therapy (3) | R-Bendamustine (5) |  |
|  | Autograft (1) | Pembrolizumab (1) | R-Ibrutinib (2) |  |
|  |  | R-Bendamustine Polatuzumab (1) | R-CHOP (1) |  |
|  |  | R-CHOP (13) | R-CVP (3) |  |
|  |  | Rituximab (1) | Rituximab (20) |  |
|  |  | R-Lenalidomide (2) | R-Varlilumab (3) |  |
|  |  | DA-R-EPOCH (1) | R-Lenalidomide (1) |  |
|  |  | DLI (1) | Cladribine (1) |  |
|  |  | R-GCVP (2) | CVP (1) |  |
|  |  | R-GCP (1) | DRC (1) |  |
|  |  | R-Gem-Ox (1) | FCR (3) |  |
|  |  |  | Obinutuzumab (3) |  |
|  |  |  | Obinutuzumab- Chlorambucil (1) |  |
|  |  |  | Venetoclax (1) |  |
| **Previous COVID-19** | 0 | 3 | 3 | 0 |

**Table S1. Baseline Characteristics of Participants.**

Table 1 shows the number of cases within each diagnostic category, the mean age and range of participants, gender, type of vaccine received for the first dose and the disease subtypes (diffuse large B-cell lymphoma (DLBCL), Epstein Barr virus (EBV), primary central nervous system lymphoma (PCNSL), follicular lymphoma (FL), chronic lymphocytic leukemia (CLL), mantle cell lymphoma (MCL), marginal zone lymphoma (MZL), enteropathy associated T-cell lymphoma (EATL), angioimmunoblastic T-cell lymphoma (AITL), breast implant associated anaplastic large cell lymphoma (BIA-ALCL)). For the treatment status, 'none' indicates participants who have never received systemic anti-lymphoma therapy, 'previous' indicates participants who completed treatment >6 months prior to the first vaccine dose, and 'active' are patients who are concurrently receiving systemic anti-lymphoma therapy at the time of administration of the first dose of vaccine, or completed treatment ≤6 months prior to the first vaccine dose, or commenced anti-systemic anti-lymphoma therapy <4 weeks after receiving the first vaccine dose. The number of lines of systemic anti-lymphoma treatment is shown, and the number of who participants who have previously received an autologous or allogeneic stem cell transplant. Their remission status with respect to their lymphoma at the time of receiving the first dose of vaccination is shown (complete remission (CR), partial remission (PR), progressive disease (PD), stable disease (SD)). The most recent lines of therapy received by participants are described (ABVD/AVD: doxorubicin, bleomycin, vinblastine, dacarbazine; ChlVPP/EVA-doxorubicin, etoposide, prednisolone, procarbazine, vinblastine, vincristine; BEACOPDac: bleomycin, etoposide, doxorubicin, cyclophosphamide, vincristine, prednisolone, dacarbazine; CAR-T therapy: chimeric antigen receptor T-cell therapy; R: rituximab; CHOP: cyclophosphamide, doxorubicin, vincristine, prednisolone; CVP: cyclophosphamide, vincristine, prednisolone; DRC: dexamethasone, rituximab, cyclophosphamide; FCR: fludarabine, cycophosphamide, rituximab). Number of participants with a history of previous COVID-19 infection, either confirmed or clinically suspected is shown.

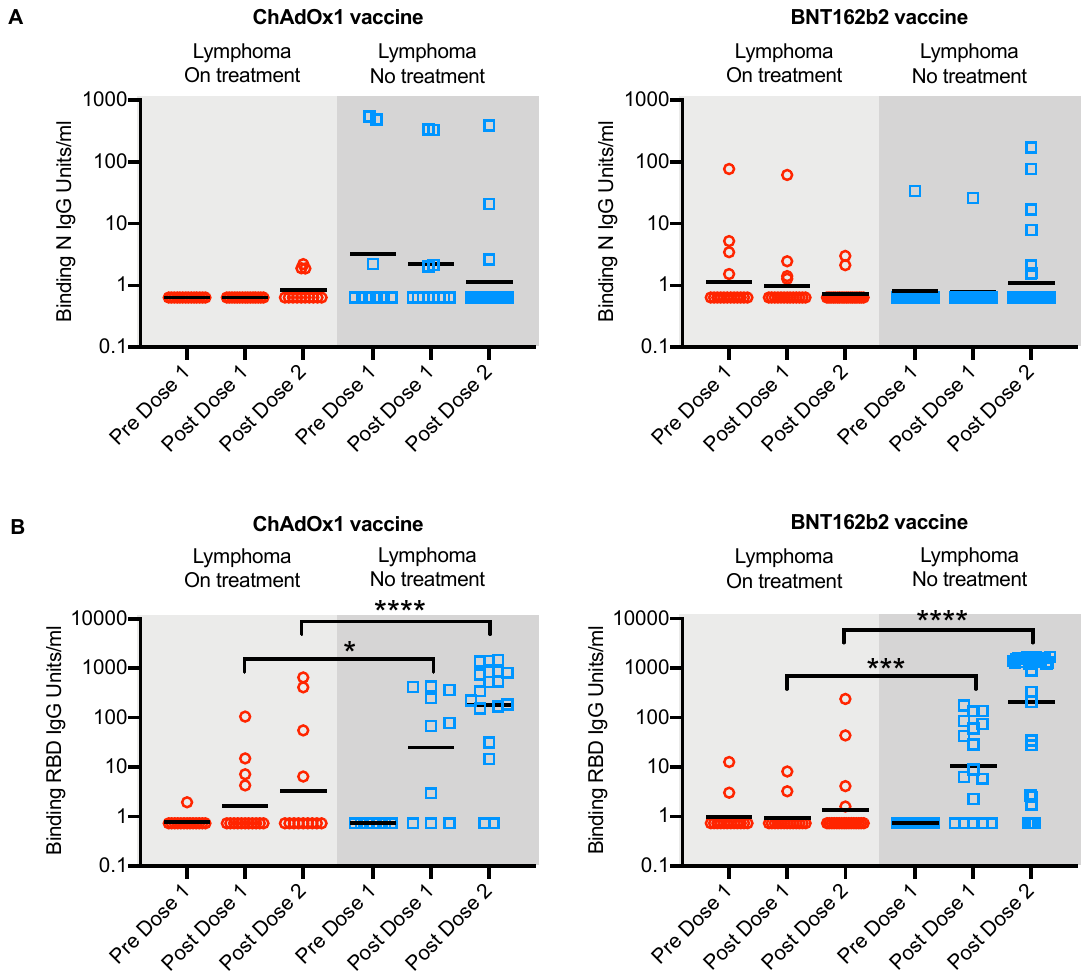

**Figure S1. IgG Responses to First and Second Doses of SARS-Cov-2 Vaccine in Lymphoma**

**(A)** IgG antibodies against SARS-CoV-2 nucleocapsid (N) antigen were measured using MSD and calibrated against the WHO COVID-19 International Reference Serum prior to vaccination (Pre Dose 1), 4 weeks post first vaccination (Post Dose 1) and/or 2-4 weeks post second vaccination (Post Dose 2) with ChAdOx1 or BNT162b2 vaccines. Red circles represent participants with lymphoma in the "on treatment" group, and blue squares represent participants with lymphoma in the "no treatment" group. Anti-N IgG ≤0.64 BAU/ml are below the lower limit of detection and anti-N >6.6 BAU/ml are considered to have had previous contact with SARS-Cov-2. These individuals are excluded from anti-RBD and anti-S IgG analysis. GMT are shown.

**(B)** As (A) but IgG antibodies against SARS-CoV-2 Receptor Binding Domain (RBD) antigen were measured. Anti-RBD IgG ≤0.73 BAU/ml are below the lower limit of detection. These individuals are excluded from anti-RBD and anti-S IgG analysis. GMT are shown. Significance was determine using two-tailed Mann-Whitney test, p<0.05(*), p<0.01(**), p<0.001(***), p<0.0001(****).
